# Clinical outcomes after viraemia among people receiving dolutegravir versus efavirenz-based first-line antiretroviral therapy in South Africa

**DOI:** 10.1101/2023.08.15.23293965

**Authors:** Kwabena Asare, Lara Lewis, Johan van der Molen, Yukteshwar Sookrajh, Thokozani Khubone, Pravikrishnen Moodley, Richard J Lessells, Kogieleum Naidoo, Phelelani Sosibo, Nigel Garrett, Jienchi Dorward

**Author notes:** **Corresponding author:** Mr Kwabena Asare, MSc. Centre for the AIDS Programme of Research in South Africa. Doris Duke Medical Research Institute, Nelson R Mandela School of Medicine, University of KwaZulu-Natal, Private Bag X7, Congella, 4013, South Africa.

## Abstract

**Introduction:** We aimed to compare clinical outcomes after viraemia between dolutegravir versus efavirenz-based first-line antiretroviral therapy (ART) as evidence is lacking outside clinical trials in resource-limited settings.

**Methods:** We conducted a retrospective cohort analysis with routine data from 59 South African clinics. We included people living with HIV ≥15 years old receiving first-line tenofovir disoproxil fumarate, lamivudine, dolutegravir (TLD) or tenofovir disoproxil fumarate, emtricitabine, efavirenz (TEE), and with first viraemia (≥50 copies/ml) between June to November 2020. We used multivariable modified Poisson regression models to compare retention-in-care and viral suppression (<50 copies/ml) after 12 months between participants on TLD versus TEE.

**Results:** At first viraemia, among 9657 participants, 6457 (66.9%) were female, median age was 37 years (interquartile range [IQR] 31-44); 7598 (78.7%) were receiving TEE and 2059 (21.3%) TLD. Retention-in-care was higher in the TLD group (84.9%) than TEE (80.8%), adjusted risk ratio (aRR) 1.03, 95%CI 1.00-1.06. Of 6569 participants retained in care and had a 12-month viral load, viral suppression was similar between TLD (78.9%) and TEE (78.8%) groups, aRR 1.02, 95%CI 0.98-1.05. However, 3368 participants changed ART during follow-up; the majority from TEE to first-line TLD (89.1%) or second-line (TLD 3.4%, zidovudine/emtricitabine/lopinavir-ritonavir 2.1%). In sensitivity analysis among the remaining 3980 participants who did not change ART during follow-up and had a 12-month viral load, viral suppression was higher in the TLD group (78.9%) than TEE (74.9%), aRR 1.07, 95%CI 1.03-1.12.

**Conclusions:** Among people with viraemia on first-line ART, dolutegravir was associated with better retention-in-care and similar or better viral suppression than efavirenz.

## INTRODUCTION

In South Africa, about 23% of people living with HIV (PLHIV) experience an episode of viraemia during first-line antiretroviral therapy (ART)[1]. Persistent viraemia during ART in PLHIV is usually due to inconsistent adherence[2, 3] or drug resistance[3-5] and increases the risk of virologic or treatment failure[6, 7]. This often leads to a slower immune reconstitution[8] and a higher incidence of all-cause mortality[9-11].

Dolutegravir is an integrase strand transfer inhibitor (INSTI) being rolled out for ART in South Africa since 2019[12] and in most low- and middle-income countries (LMICs)[13], replacing the previous drug efavirenz. Compared to efavirenz and other non-nucleoside reverse transcriptase (NNRTI) based regimens, dolutegravir is more effective and tolerable with an increased genetic barrier against drug resistance based on clinical trial evidence[14-18]. Accordingly, people receiving first-line dolutegravir-based regimens who present with viraemia may be more likely to have inconsistent treatment adherence rather than drug resistance and could be more likely to virally re-suppress without the need for changing regimens, unlike people receiving efavirenz[19].

Based on these therapeutic strengths of dolutegravir, the World Health Organisation 2021 ART treatment guidelines recommend delaying switching to second-line ART in people with virological failure on first-line dolutegravir-based regimens[20]. This contrasts with recommendations for an early switch to second-line ART among people with virological failure receiving first-line efavirenz and other NNRTI-based regimens[20]. However, there is limited evidence from routine healthcare settings on clinical outcomes and viral load trajectories after viraemia in people receiving first-line dolutegravir-based ART from high HIV prevalence settings in LMICs.

Therefore, we aimed to assess retention-in-care and viral load trajectories after viraemia in people receiving first-line dolutegravir-based ART compared to those receiving efavirenz.

## METHODS

### Study design and setting

We conducted a retrospective cohort study with de-identified, routinely collected data from 59 public, primary healthcare facilities in eThekwini Municipality, KwaZulu-Natal, South Africa. In these clinics, viral load testing is done at 6 and 12 months after ART initiation and then 12-monthly thereafter[12]. CD4 count is routinely measured at ART initiation and after 12 months and subsequently repeated if clinically indicated (e.g., viral load ≥1000 copies/ml).

The 2019 South African HIV treatment guideline[12] recommends that PLHIV with a viral load ≥50 copies/ml during first-line ART should receive enhanced adherence counselling, and a repeat viral load should be performed after two to three months. People receiving first-line ART with two consecutive viral loads ≥1000 copies/ml two to three months apart are classified as having virological failure, and switching to second-line ART is recommended if they were receiving an NNRTI-based regimen including efavirenz or nevirapine. However, for those receiving dolutegravir, switching to second-line ART is only recommended after two years of ongoing viraemia.

This study was approved by the Biomedical Research Ethics Committee of the University of Kwazulu-Natal (BE646/17), the KwaZulu-Natal Provincial Health Research Ethics Committee (KZ_201807_021), the TB/HIV Information Systems Data Request Committee, and the eThekwini Municipality Health Unit.

### Data sources and data management

The data source for this study was South Africa’s TIER.Net electronic database which contains demographics, clinical status, regimen and clinic visit information of people receiving ART in public sector healthcare clinics[21]. All datasets were de-identified by the South African National Department of Health’s TB/HIV Information Systems (THIS-www.tbhivinfosys.org.za/) before access and use. We performed data cleaning to remove duplicate entries and rationalize ART regimen lines according to clinical guidelines.

### Participants

The study cohort included PLHIV aged ≥15 years with first viraemia (viral load ≥50 copies/ml) between June 01, 2020, and November 30, 2020, while receiving first-line TEE or TLD regimens. The TLD group included participants who initiated TLD and who transitioned from an NNRTI-based regimen to TLD before viraemia. The ART regimen lines were based on a predefined variable from the TIER.Net dataset, prevailing guidelines, and clinical considerations. For example, someone who transitioned from TEE to TLD with two previous viral loads ≥1000 copies/ml was reclassified as second-line TLD. The ART regimen at viraemia was defined as the regimen participants were receiving at the time of viraemia, so all participants who transitioned from TEE to TLD on the date of viraemia were classified as being on TEE at the time of viraemia. We selected the baseline period of viraemia to include as many participants as possible on dolutegravir as implementation started in December 2019 and to allow a minimum of 365 days (12 months) plus 90 days of follow-up before the data cutoff of our dataset on April 30, 2022. We excluded participants who had been receiving their ART regimen at the time of viraemia for less than 90 days and those not receiving standard first-line regimens of TEE or TLD at the time of viraemia.

### Primary outcomes

Our primary outcomes were retention-in-care and viral suppression at 12 months follow-up after viraemia. Retention-in-care at 12 months was defined as not being lost to follow-up or recorded in TIER.Net as either deceased or ‘transferred out’ to another clinic (as we could not access or link to data at other clinics to establish subsequent retention-in-care) by 365 days after viraemia. Loss to follow-up was defined based on the South African ART programme guidelines of being ≥90 days late for a visit[22]. Viral suppression was defined as viral load <50 copies/ml. Considering viral loads are not always completed on time in routine care, we defined the 12-month window as the closest viral load to 365 days between 181 to 545 days after viraemia. We included only the viral loads of participants retained in care.

### Secondary outcomes

To assess implementation fidelity to the guidelines for managing viraemia, we evaluated the secondary outcomes of clinic visit attendance up to 6 months after viraemia and 3-month repeat viral load completion and viral suppression (<50 copies/ml). We defined the 3-month viral load window as the closest viral load testing date to 90 days between 28 to 180 days after viraemia.

### Exposures

The primary exposure was the first-line ART regimen combination (TLD versus TEE) that participants were receiving at the time of viraemia. Potential confounders included participant characteristics at viraemia, including age, gender, active tuberculosis disease, viral load category, CD4 count category, and time-period of viraemia.

### Statistical analyses

We performed all statistical analyses using R 4.2.0 (R Foundation for Statistical Computing, Vienna, Austria)[23]. We used modified Poisson regression models with robust standard errors adjusting for clustering by clinic[24] to determine the risk ratios of retention-in-care at 12 months and viral suppression at 3 and 12 months after viraemia. In all regression models, we compared TLD versus TEE first-line regimens and adjusted for participant characteristics at viraemia: age, gender, active tuberculosis disease, time-period of viraemia, viral load category and CD4 count category.

We performed two separate sensitivity analyses. The first sensitivity analysis involved the 12-month retention-in-care outcome, where we included participants who were transferred out to another clinic as being retained in care. The second sensitivity analysis involved the 12-month viral suppression outcome, where we excluded participants who changed their ART regimen during the 12-month follow-up.

To better understand management and outcomes among people with high-level viraemia, we conducted a secondary analysis in participants with viraemia ≥1000 copies/ml. In this sub-group, we plotted a Sankey diagram to graphically present viral load trajectories after viraemia and switching to second-line ART (only in the TEE group). We only included participants with complete 3-and 12-month viral load results for the Sankey diagram.

## RESULTS

### Cohort characteristics at the time of viraemia

Between June 01, 2020, and November 30, 2020, 11366 people aged ≥15 had viraemia while receiving first-line ART at the study clinics (Figure 1). At the time of viraemia, 1389 had been receiving their current regimen for less than 90 days, and 320 participants were not receiving standard TEE or TLD regimens and were excluded. Of the remaining 9657 people included in the analyses, 7598 (78.7%) were receiving TEE, and 2059 (21.3%) were receiving TLD regimens at the time of viraemia (Table 1). In the TLD group, 584 (28.4%) had been initiated on TLD, and 1475 (71.6%) had transitioned to TLD from an NNRTI-based regimen before viraemia. The time on the current regimen was lower in the TLD group (0.5 years, IQR 0.4-0.5) than in the TEE group (3.0 years, IQR 1.1-5.3). The median age of the cohort was 37 years (IQR 31-44), and 6457 (66.9%) were female, of whom 196 (3.0%) were pregnant. There were more females (n=5624, 74.0%) in the TEE group and more males (n=1226, 59.5%) in the TLD group.

**Table 1.**
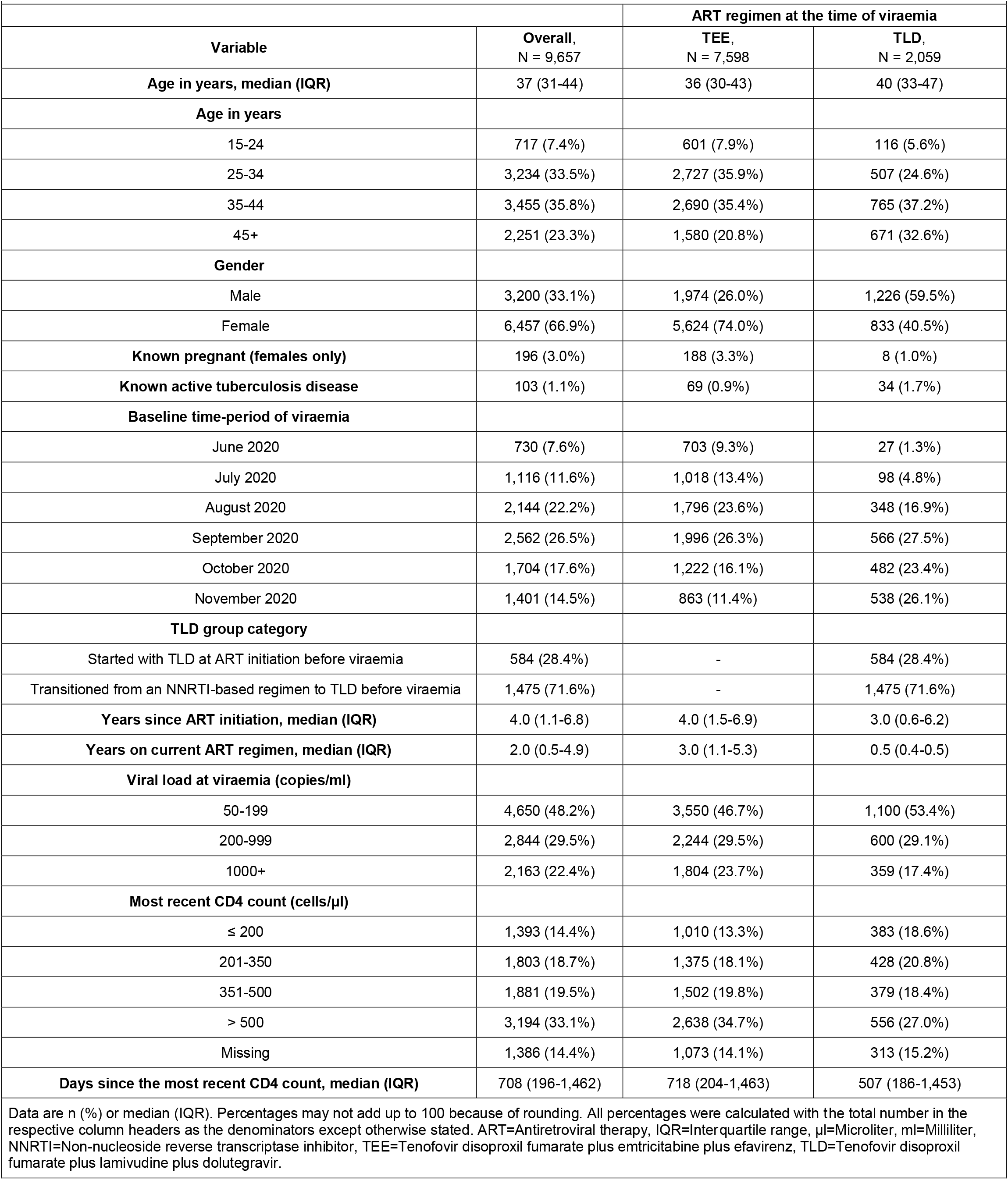
Baseline characteristics of people with viraemia (≥50 copies/ml) while receiving first-line ART.

**Figure 1.**
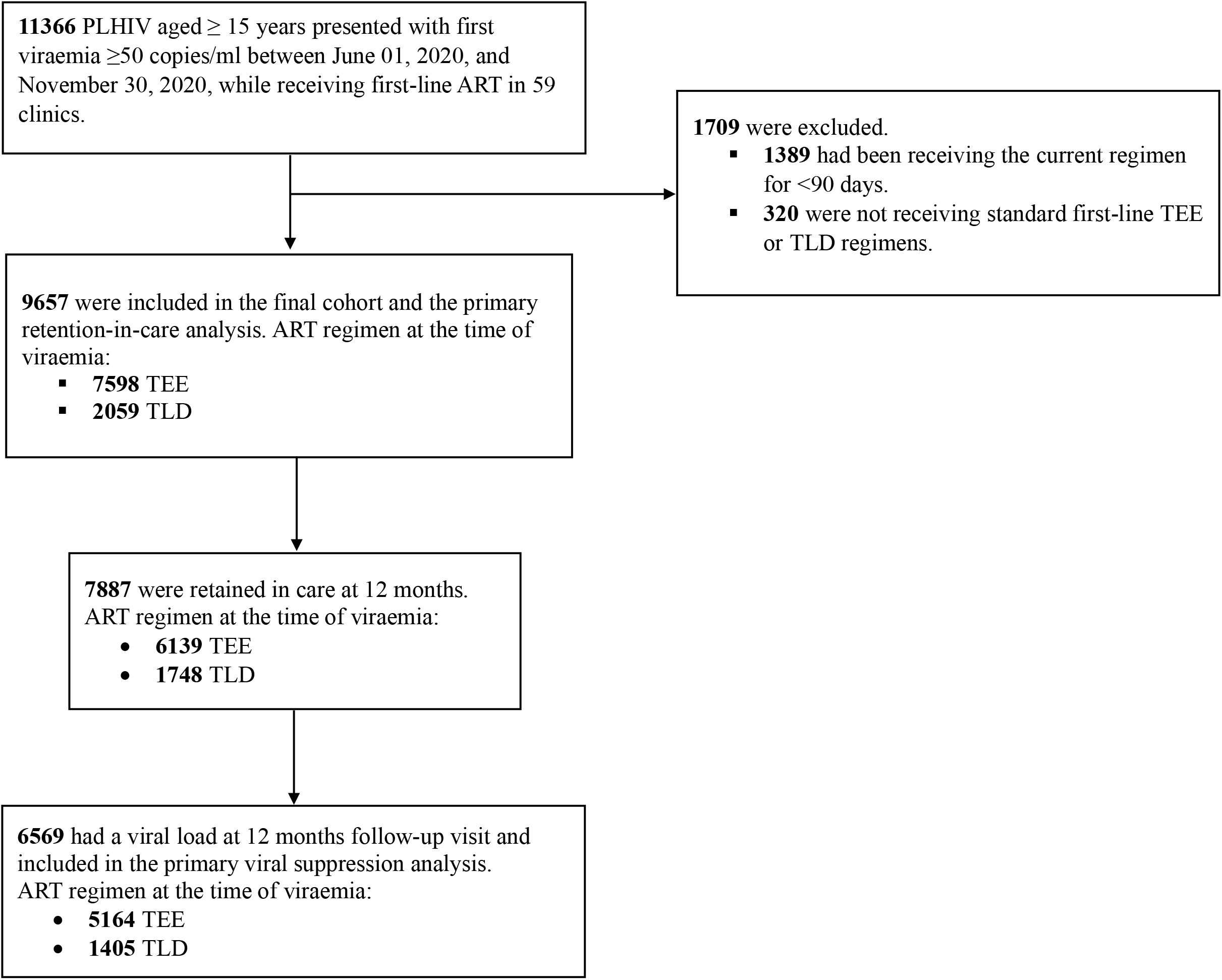
Flow diagram of PLHIV receiving antiretroviral therapy at 59 clinics in South Africa. ART=Antiretroviral therapy, ml=Milliliter, PLHIV=People living with HIV, TEE=Tenofovir disoproxil fumarate plus emtricitabine plus efavirenz, TLD=Tenofovir disoproxil fumarate plus lamivudine plus dolutegravir.

### Clinical outcomes after viraemia

Twelve months after viraemia, 1183 (12.3%) participants were recorded as lost to follow-up, 59 (0.6%) had died, 528 (5.5%) had transferred out to another clinic, and 7887 (81.7%) were retained in care (Table 2). Retention-in-care at 12 months was 80.8% (n=6139) in the TEE group and 84.9% (n=1748) in the TLD group (Table 2). In the multivariable Poisson regression analysis adjusted for age, gender, active tuberculosis disease, time-period of viraemia, viral load category and CD4 count category, all at the time of viraemia, 12-month retention-in-care was higher in the TLD group than TEE (adjusted risk ratio [aRR] 1.03, 95% confidence interval [CI] 1.00-1.06, p=0.047) (Table 3). In sensitivity analysis (Table S 1), where we classified people who were transferred out to another clinic as being retained in care, 12-month retention-in-care was similar between the TLD (89.1%) and TEE (86.6%) groups, aRR 1.02, 95% CI 0.99-1.04, p=0.149.

**Table 2.**
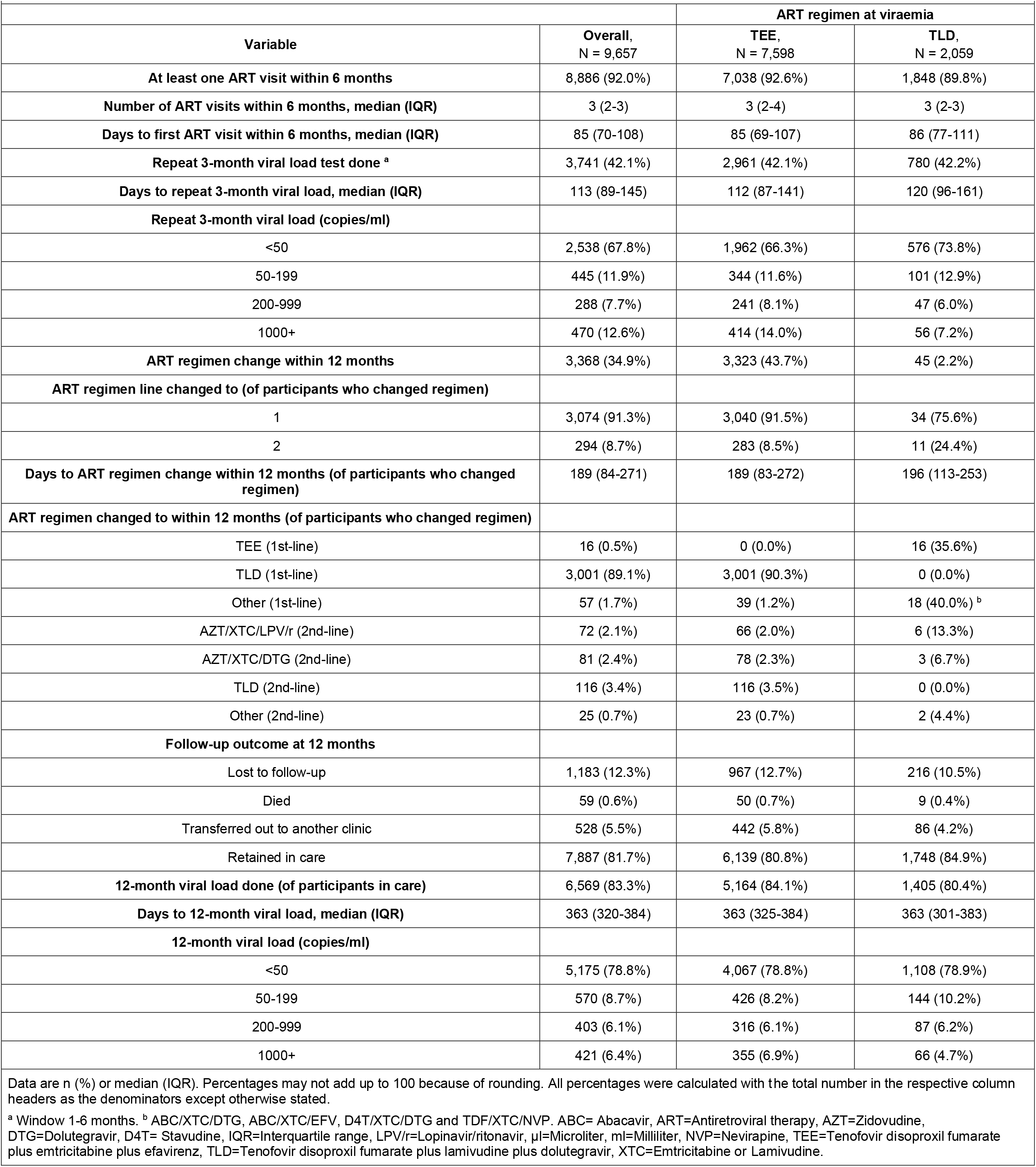
Follow-up outcomes after viraemia (≥50 copies/ml) in people receiving first-line ART.

**Table 3.**
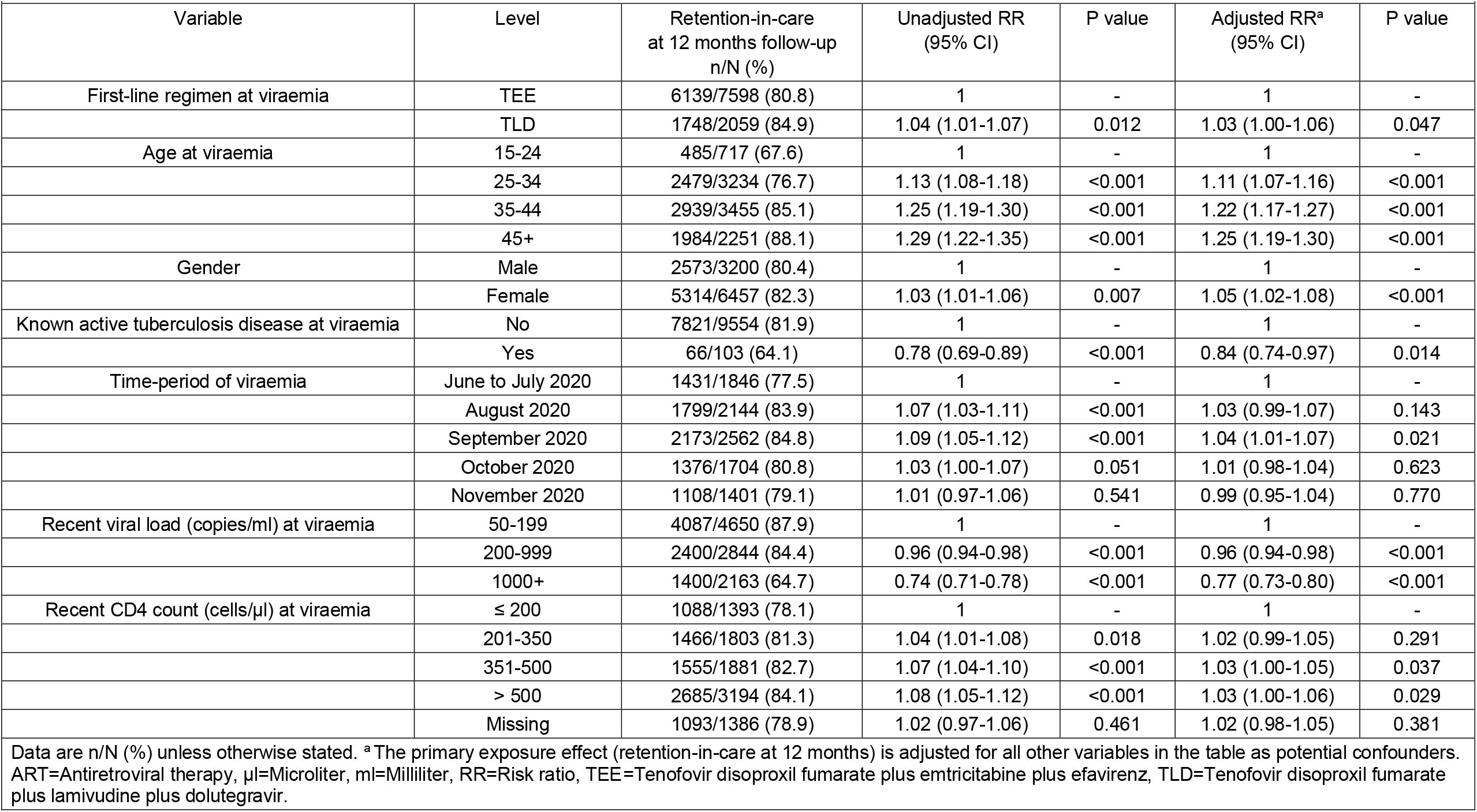
Univariable and multivariable Poisson regression models of factors associated with retention-in-care at 12 months follow-up after viraemia (≥50 copies/ml) in people receiving first-line ART (N=9657)

Of participants retained in care at 12 months, 6569 (83.3%) had a follow-up viral load done at a median of 363 days (IQR, 320-384) after viraemia. By regimen, 5164 (84.1%) in the TEE group and 1405 (80.4%) in the TLD group had a 12-month viral load (Table 2). Of participants with a 12-month viral load, 4067 (78.8%) in the TEE group and 1108 (78.9%) in the TLD group were virally suppressed. The multivariable Poisson regression analysis showed no difference in 12-month viral suppression in the TLD group versus TEE (aRR 1.02, 95% CI 0.98-1.05, p=0.418) (Table 4).

**Table 4.**
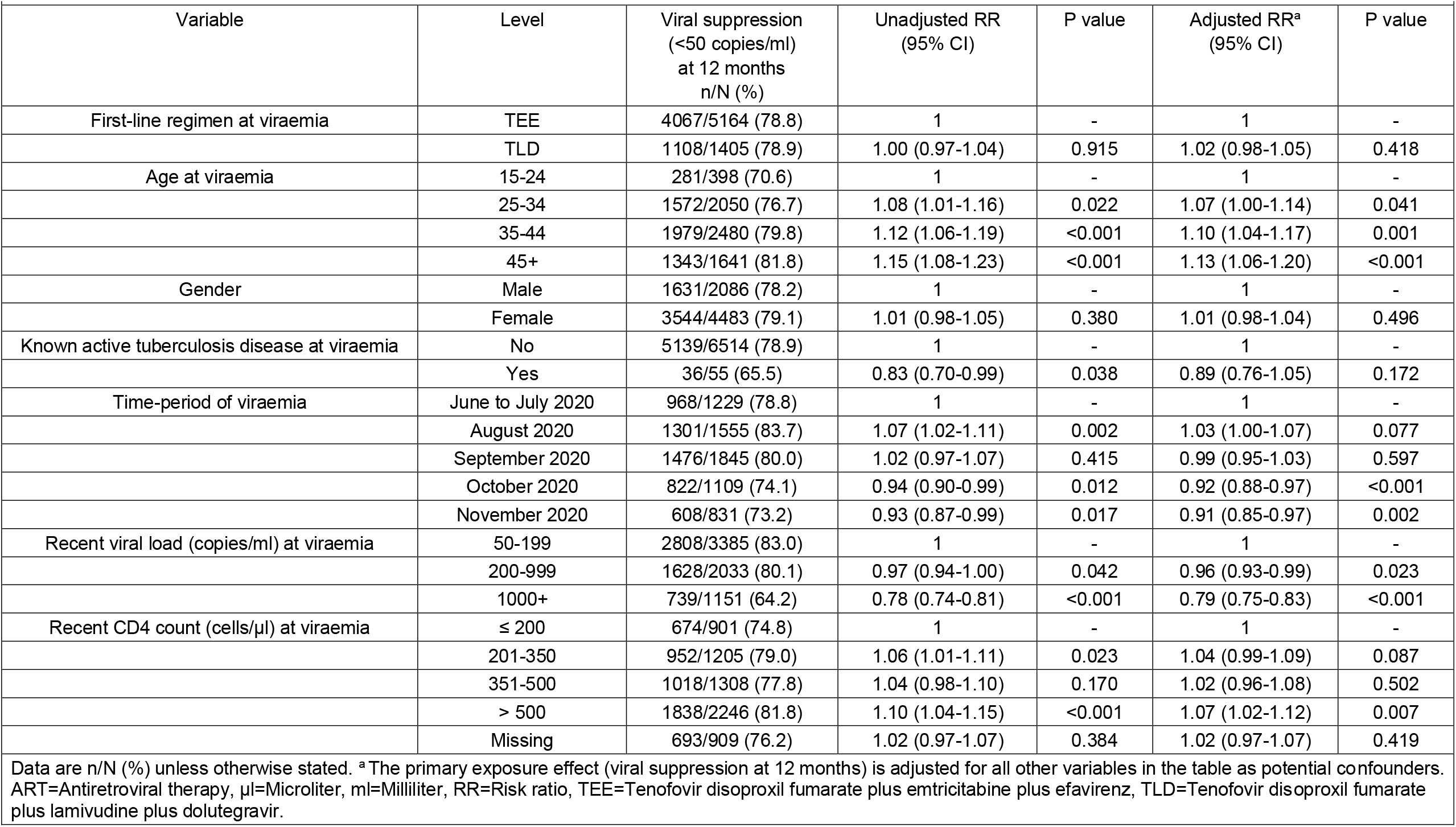
Univariable and multivariable Poisson regression models of factors associated with viral suppression at 12 months follow-up after viraemia (≥50 copies/ml) in people receiving first-line ART who were retained-in-care at 12 months follow-up and had viral load done (N=6569)

However, at a median of 189 days (IQR, 84-271) after viraemia, 3368 (34.9%) participants changed their ART regimen (Table 2). Of these, 3074 (91.3%) transitioned to another first-line, and 294 (8.7%) switched to second-line regimens. Participants in the TEE group had more regimen changes after viraemia (n=3323, 43.7%) than those in the TLD group (n=45, 2.2%). In a sensitivity analysis among 3980 participants who did not change their regimen within 12 months after viraemia and had a 12-month viral load, 12-month viral suppression was more likely in the TLD group (78.9%) than TEE (74.9%), aRR 1.07, 95% CI 1.03-1.12, p=0.001) (Table 5).

**Table 5.**
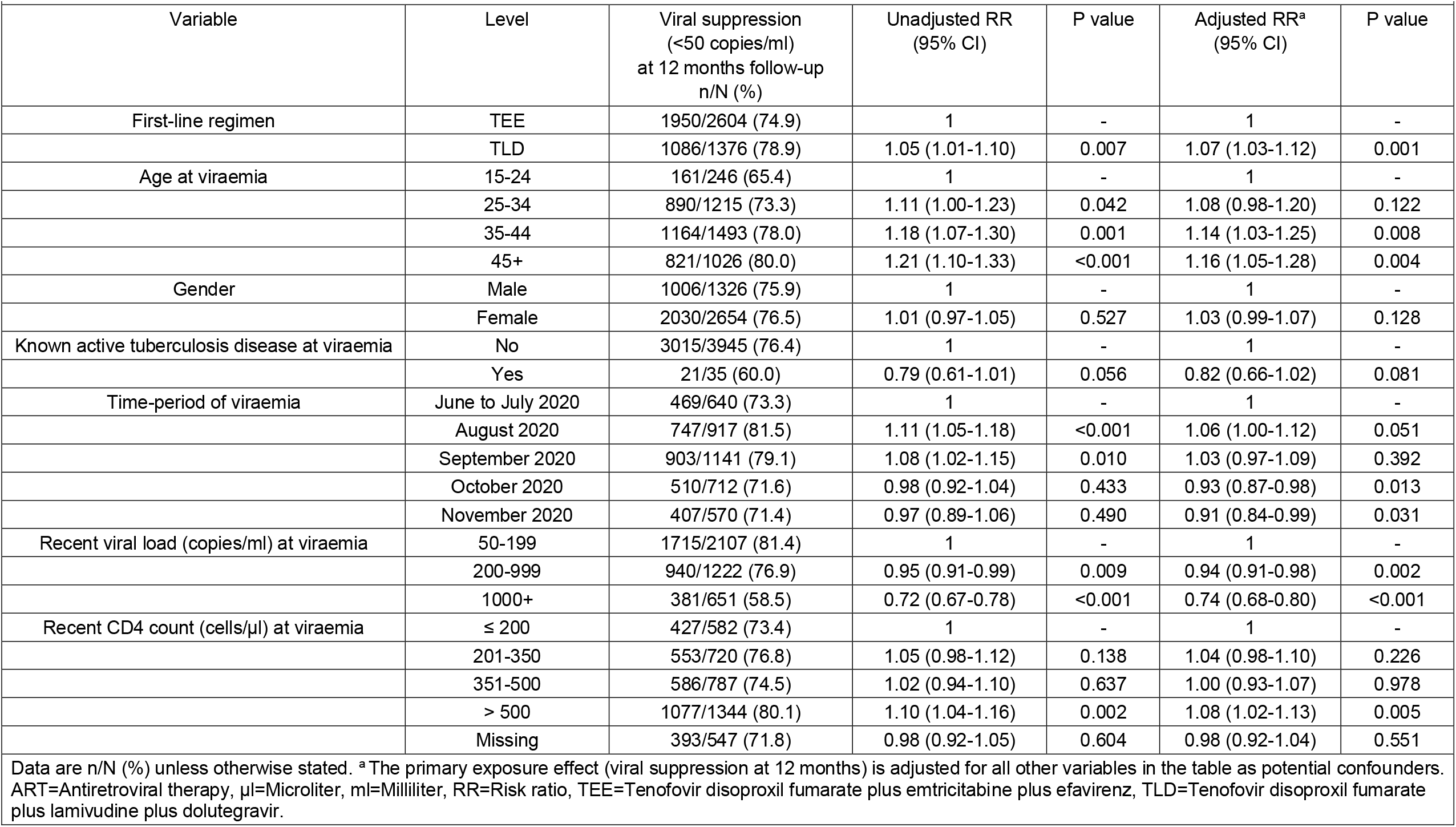
Sensitivity analysis: Univariable and multivariable Poisson regression models of factors associated with viral suppression at 12 months follow-up after viraemia (≥50 copies/ml) in people receiving first-line ART who did not change their ART regimen within 12 months, were retained in care at 12 months follow-up and had viral load done (N=3980)

We assessed implementation fidelity to viraemia management guidelines. In the first 6 months after viraemia, 7038 (92.6%) participants in the TEE group and 1848 (89.8%) in the TLD group had at least one visit after a median of 85 (IQR, 69-107) and 86 (IQR, 77-111) days respectively. Within the 6 months, participants in the TEE group had a median of 3 (IQR, 2-4) visits, and participants in the TLD group had a median of 3 (IQR, 2-3) visits (Table 2). Regarding viral load monitoring after viraemia, only 2961 (42.1%) participants in the TEE group and 780 (42.2%) in the TLD group had a 3-month repeat viral load done after a median of 112 (IQR, 87-141) and 120 (IQR, 96-161) days respectively. Three-month repeat viral load suppression was higher in the TLD group (n=576,73.8%) than the TEE group (n=1962, 66.3%), aRR 1.07, 95% CI 1.02-1.13, p=0.009; Table S 2).

We also assessed follow-up outcomes restricted to participants with high-level viraemia (≥1000 copies/ml). A total of 2163 participants presented with high-level viraemia, of whom 1804 83.4%%) were receiving TEE and 359 (16.6%) were receiving TLD (Table S 3). Of these, 328 (34.7%) participants in the TEE group and 38 (20.0%) in the TLD group had a virological failure (repeat 3-month viral load ≥1000 copies/ml). Of participants with virological failure in the TEE group, 104 (31.7%) were switched to second-line ART within the 12-month follow-up period (from the time of viraemia). In the multivariable Poisson regression models, participants who were on TLD at the time of high-level viraemia had better outcomes than those on TEE regarding 3-month viral suppression (63.7% versus 42.3%, aRR 1.39, 95% CI 1.22-1.58, p<0.001, Table S 4), 12-month retention-in-care (71.9% versus 63.3%, aRR 1.11, 95% CI 1.02-1.22, p=0.018, Table S 5) and 12-month viral suppression (76.6% versus 61.6%, aRR 1.24, 95% CI 1.12-1.36, p<0.001, Table S 6). A graphical overview of outcomes after high-level viraemia is presented in the Sankey diagram in Figure 2.

**Figure 2.**
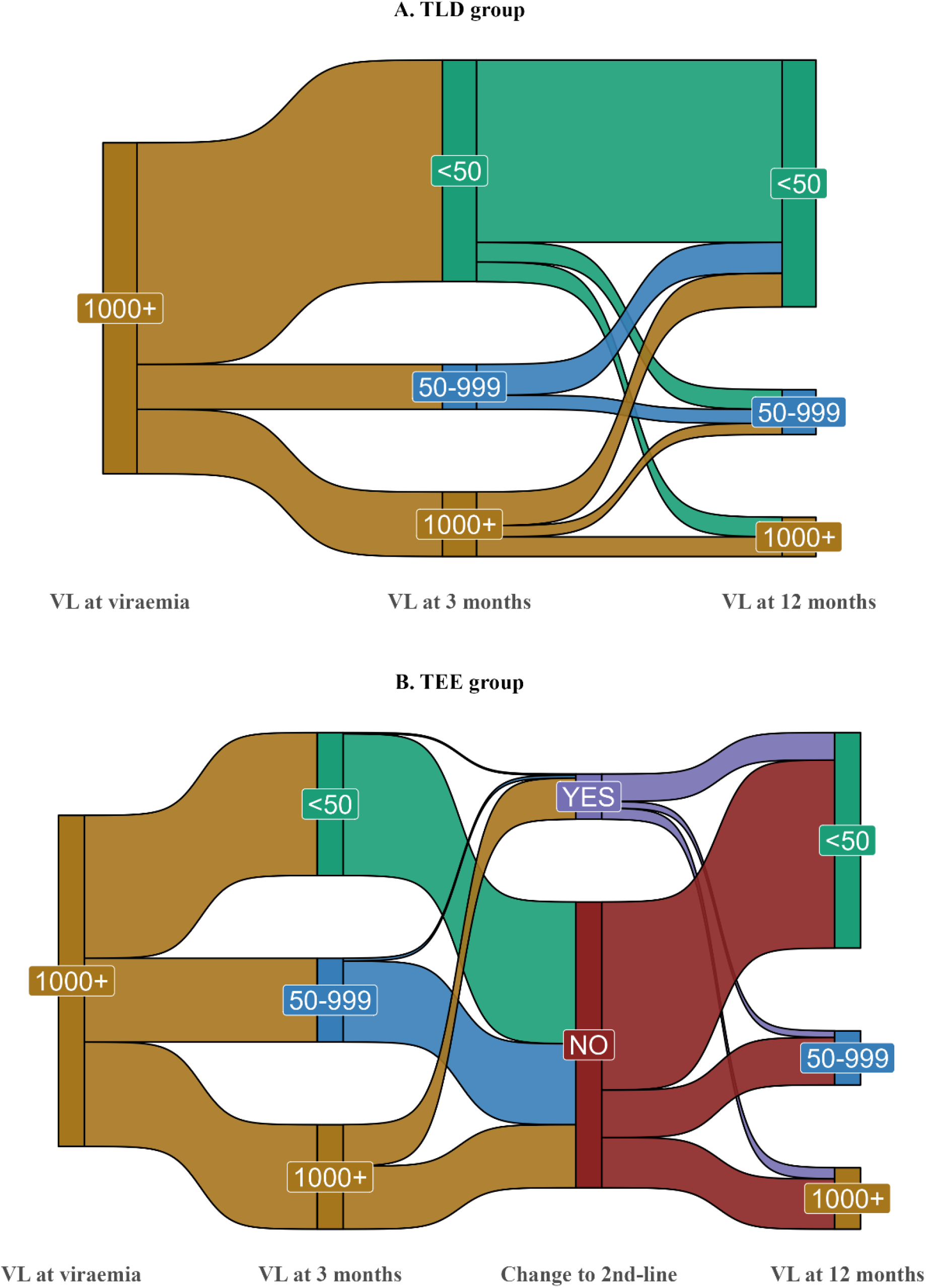
Sankey diagram showing the flow of viraemia management outcomes in PLHIV with high-level viraemia (≥1000 copies/ml) while receiving first-line antiretroviral therapy for ≥90 days at the time of viraemia. Panel (A) TLD group and panel (B) TEE group. ml=Milliliter, PLHIV=People living with HIV, TEE=Tenofovir disoproxil fumarate plus emtricitabine plus efavirenz, TLD=Tenofovir disoproxil fumarate plus lamivudine plus dolutegravir, VL=Viral load (copies/ml).

In the Poisson regression analyses, outcomes were generally less likely among participants aged <25 at the time of viraemia than older participants.

## Discussion

In this cohort study with large-scale ART programmatic data from 59 public sector healthcare clinics in South Africa, retention-in-care after viraemia was better among PLHIV on first-line TLD than TEE. Viral suppression after viraemia was better among PLHIV, who stayed on first-line TLD than those who stayed on first-line TEE. Retention-in-care and viral suppression were better among participants with high-level viraemia (≥1000 copies/ml) on TLD than TEE.

Limited evidence exists on retention-in-care and viral suppression after viraemia on first-line dolutegravir-based ART in routine LMIC settings, but such data is urgently needed to optimise strategies for managing viraemia on dolutegravir. Evidence from clinical trials has shown lower rates of treatment discontinuation and abandonment due to lower rates of adverse events with dolutegravir-based than efavirenz-based ART regimens[17, 18]. Our finding of better retention-in-care with TLD than TEE after viraemia on first-line ART is consistent with the results from these studies[17, 18]. It emphasises the potential effect of the tolerability of dolutegravir in ensuring consistent treatment adherence and improved clinical outcomes after viraemia.

For viral suppression after viraemia, we found one similar study conducted in South Africa among 385 participants enrolled in the ADVANCE trial who had viraemia while receiving first-line dolutegravir-based or efavirenz-based ART regimens[19]. The study used a protocol-defined virologic failure of either ≥1000 copies/ml at week 12, ≥200 copies/ml at week 24, or ≥50 copies/ml at week 48 after enrolment. In participants with follow-up viral load available, viral suppression (<50 copies/ml) within three follow-up visits after the first protocol-defined virologic failure was 74.0% (n=77/104) with tenofovir alafenamide (TAF)/emtricitabine (FTC)/dolutegravir, 85.0% (n=85/100) with TDF/FTC/dolutegravir and 40.4% (n=44/109) with TDF/FTC/efavirenz[19]. This evidence from the ADVANCE trial is consistent with our findings from routine healthcare settings, but the efavirenz group in the ADVANCE trial had lower viral suppression.

Our findings are essential in resource-limited settings approaching a full rollout to dolutegravir-based regimens for monitoring implementation successes. We used South African guideline definitions of viraemia, retention-in-care, and viral suppression and adjusted for potential confounders of clinical outcomes after viraemia, such as age, gender, active tuberculosis disease, time-period of viraemia, viral load and CD4 count. Our findings support 2019[12] and current 2023[25] South African ART treatment guidelines that recommend the delay of switching to second-line ART after virologic failure among participants receiving first-line dolutegravir-based regimens since enhanced adherence counselling is more likely to lead to re-suppression due to dolutegravir’s efficacy[14] and increased genetic barrier to drug resistance[26].

Furthermore, our analysis on viraemia management revealed healthcare bottlenecks that can be addressed to improve clinical outcomes after viraemia. Although about 90% of participants had a clinic visit within the first 6 months after viraemia, less than half had a repeat 3-month viral load. Some of these missing repeat viral loads might have been done but not recorded in TIER.Net. However, studies in South Africa[27] and Lesotho[28] that used prospectively collected clinic data have shown similar rates of 47.7% and 40.0% of repeat viral load completion within 6 months after the first elevated viral load ≥1000 copies/ml in participants receiving ART. These gaps lead to missed opportunities for adequate viraemia management, such as confirming persistent viraemia and switching to second-line ART with potentially negative implications for increased morbidity and mortality[29, 30].

Our analysis also revealed poorer retention-in-care and viral suppression outcomes among younger PLHIV. Younger people remain a high-risk group of the HIV epidemic[31], and these findings indicate they might also struggle to achieve better treatment outcomes during chronic HIV infection. Thus, engaging and paying particular attention to young people with viraemia on first-line ART might also be necessary as they likely have special needs that must be addressed for improved outcomes.

Our study had some limitations. Although most people currently initiating ART in South Africa are prescribed dolutegravir-based regimens[32], new initiations on dolutegravir and the transition from other regimens to dolutegravir are not random. People initiating or transitioning to dolutegravir may be more likely to be clinically stable than people on different regimens, so we adjusted for the viral load category at the time of viraemia in all regression models. However, we cannot rule out confounding by variables not recorded in the dataset. The timing of viraemia was also different as more participants in the TEE group were viraemic during the COVID-19 period and might have been at a higher risk of interruptions in care post-viraemia. However, we adjusted for the time-period of viraemia in our analyses. This study is also at the early stages of the dolutegravir implementation in South Africa, and more extended follow-up data are needed as dolutegravir resistance may increase over time. Our analysis is also limited to one province, and a national-level study would be more representative of the South African population.

In conclusion, our findings demonstrate that, among people with viraemia during first-line ART in routine healthcare settings, dolutegravir use was associated with better retention-in-care and better viral suppression among people who did not change ART regimen. Among people with high-level viraemia ≥1000 copies/mL, TLD was even more strongly associated with improved retention-in-care and viral suppression. Improving adherence to guidelines for managing viraemia, including enhanced adherence counselling and repeat viral load monitoring, is important for better outcomes with TLD after viraemia. Further research to investigate longer-term outcomes, such as switching to second-line ART after two years of ongoing viraemia on first-line dolutegravir, would be a great addition to the evidence.

## Supporting information

Supplementary results

## Data Availability

We cannot publicly share the data used for this analysis because of the legal and ethical requirements regarding the use of routinely collected clinical data in South Africa. Interested parties can request access to the data from the eThekwini Municipality Health Unit and the South African National Department of Health TB/HIV Information System (contact details obtainable upon request to JD).

## Notes

### Author contributions

KA, YS, LL, JvdM, RJL, KN, NG and JD conceptualised the study. TK, YS, PM, and PS managed data collection. TK, PS, and JvdM oversaw data curation. KA, YS, TK, JvdM, PS, LL, RvH, NG and JD had full access to the data in the study. KA, JvdM, LL, and JD analysed the data. KA drafted the manuscript. All authors reviewed and approved the final version for submission.

## Acknowledgements

We acknowledge the eThekwini Municipality Health Unit staff, patients, and primary care clinics.

