## Supplementary results for "Clinical outcomes after viraemia among people receiving dolutegravir versus efavirenz-based first-line antiretroviral therapy in South Africa"

### Supplementary tables

| **Table S 1.** Univariable and multivariable Poisson regression models of factors associated with retention-in-care at 12 months follow-up after viraemia (≥50 copies/ml) in people receiving first-line ART (classifying participants who transferred out to another clinic as retained in care) (N=9657) | | | | | | |
| --- | --- | --- | --- | --- | --- | --- |
| Variable | Level | Retention-in-care  at 12 months  n/N (%) | Unadjusted RR  (95% CI) | P value | Adjusted RR^a^  (95% CI) | P value |
| First-line regimen at viraemia | TEE | 6581/7598 (86.6) | 1 | - | 1 | - |
|  | TLD | 1834/2059 (89.1) | 1.02 (1.00-1.04) | 0.105 | 1.02 (0.99-1.04) | 0.149 |
| Age at viraemia | 15-24 | 553/717 (77.1) | 1 | - | 1 | - |
|  | 25-34 | 2711/3234 (83.8) | 1.08 (1.04-1.12) | <0.001 | 1.07 (1.04-1.11) | <0.001 |
|  | 35-44 | 3080/3455 (89.1) | 1.14 (1.11-1.19) | <0.001 | 1.13 (1.09-1.16) | <0.001 |
|  | 45+ | 2071/2251 (92.0) | 1.18 (1.13-1.23) | <0.001 | 1.16 (1.11-1.20) | <0.001 |
| Gender | Male | 2716/3200 (84.9) | 1 | - | 1 | - |
|  | Female | 5699/6457 (88.3) | 1.04 (1.03-1.06) | <0.001 | 1.05 (1.03-1.08) | <0.001 |
| Known active tuberculosis disease at viraemia | No | 8340/9554 (87.3) | 1 | - | 1 | - |
|  | Yes | 75/103 (72.8) | 0.84 (0.75-0.93) | 0.002 | 0.89 (0.80-0.99) | 0.038 |
| Time-period of viraemia | June to July 2020 | 1540/1846 (83.4) | 1 | - | 1 | - |
|  | August 2020 | 1913/2144 (89.2) | 1.06 (1.03-1.09) | <0.001 | 1.03 (1.00-1.06) | 0.056 |
|  | September 2020 | 2305/2562 (90.0) | 1.07 (1.04-1.10) | <0.001 | 1.04 (1.01-1.06) | 0.008 |
|  | October 2020 | 1462/1704 (85.8) | 1.02 (0.99-1.05) | 0.114 | 1.00 (0.98-1.03) | 0.739 |
|  | November 2020 | 1195/1401 (85.3) | 1.02 (0.98-1.05) | 0.327 | 1.00 (0.97-1.03) | 0.887 |
| Recent viral load (copies/ml) at viraemia | 50-199 | 4295/4650 (92.4) | 1 | - | 1 | - |
|  | 200-999 | 2548/2844 (89.6) | 0.97 (0.96-0.99) | <0.001 | 0.97 (0.96-0.99) | <0.001 |
|  | 1000+ | 1572/2163 (72.7) | 0.79 (0.76-0.83) | <0.001 | 0.81 (0.78-0.84) | <0.001 |
| Recent CD4 count (cells/μl) at viraemia | ≤ 200 | 1174/1393 (84.3) | 1 | - | 1 | - |
|  | 201-350 | 1557/1803 (86.4) | 1.03 (1.00-1.05) | 0.058 | 1.00 (0.98-1.03) | 0.691 |
|  | 351-500 | 1654/1881 (87.9) | 1.05 (1.02-1.08) | <0.001 | 1.02 (0.99-1.04) | 0.195 |
|  | > 500 | 2846/3194 (89.1) | 1.06 (1.03-1.10) | <0.001 | 1.02 (0.99-1.04) | 0.139 |
|  | Missing | 1184/1386 (85.4) | 1.02 (0.99-1.05) | 0.232 | 1.01 (0.99-1.04) | 0.281 |
| Data are n/N (%) unless otherwise stated. ^a^ The primary exposure effect (retention-in-care at 12 months) is adjusted for all other variables in the table as potential confounders. ART=Antiretroviral therapy, μl=Microliter, ml=Milliliter, RR=Risk ratio, TEE=Tenofovir disoproxil fumarate plus emtricitabine plus efavirenz, TLD=Tenofovir disoproxil fumarate plus lamivudine plus dolutegravir. | | | | | | |

| **Table S 2.** Univariable and multivariable Poisson regression models of factors associated with 3-month viral suppression after viraemia (≥50 copies/ml) in people receiving first-line ART (N=3741) | | | | | | |
| --- | --- | --- | --- | --- | --- | --- |
| Variable | Level | 3-month viral suppression  (<50 copies/ml)  n/N (%) | Unadjusted RR  (95% CI) | P value | Adjusted RR  (95% CI) | P value |
| First-line regimen at viraemia | TEE | 1962/2961 (66.3) | 1 | - | 1 | - |
|  | TLD | 576/780 (73.8) | 1.11 (1.06-1.17) | <0.001 | 1.07 (1.02-1.13) | 0.009 |
| Age at viraemia | 15-24 | 188/316 (59.5) | 1 | - | 1 | - |
|  | 25-34 | 822/1270 (64.7) | 1.07 (0.94-1.22) | 0.287 | 1.08 (0.96-1.21) | 0.199 |
|  | 35-44 | 940/1335 (70.4) | 1.16 (1.05-1.28) | 0.004 | 1.14 (1.04-1.25) | 0.006 |
|  | 45+ | 588/820 (71.7) | 1.17 (1.06-1.29) | 0.002 | 1.14 (1.04-1.25) | 0.006 |
| Gender | Male | 826/1251 (66.0) | 1 | - | 1 | - |
|  | Female | 1712/2490 (68.8) | 1.04 (1.00-1.09) | 0.054 | 1.04 (0.99-1.09) | 0.132 |
| Known active tuberculosis disease at viraemia | No | 2522/3701 (68.1) | 1 | - | 1 | - |
|  | Yes | 16/40 (40.0) | 0.58 (0.40-0.84) | 0.004 | 0.70 (0.50-1.00) | 0.049 |
| Time-period of viraemia | June to July 2020 | 351/671 (52.3) | 1 | - | 1 | - |
|  | August 2020 | 599/830 (72.2) | 1.36 (1.25-1.49) | <0.001 | 1.24 (1.16-1.33) | <0.001 |
|  | September 2020 | 733/1005 (72.9) | 1.38 (1.26-1.50) | <0.001 | 1.27 (1.18-1.36) | <0.001 |
|  | October 2020 | 460/653 (70.4) | 1.33 (1.23-1.43) | <0.001 | 1.26 (1.17-1.35) | <0.001 |
|  | November 2020 | 395/582 (67.9) | 1.29 (1.21-1.38) | <0.001 | 1.22 (1.14-1.31) | <0.001 |
| Recent viral load (copies/ml) at viraemia | 50-199 | 1117/1377 (81.1) | 1 | - | 1 | - |
|  | 200-999 | 900/1229 (73.2) | 0.90 (0.85-0.94) | <0.001 | 0.89 (0.85-0.94) | <0.001 |
|  | 1000+ | 521/1135 (45.9) | 0.57 (0.53-0.61) | <0.001 | 0.59 (0.55-0.64) | <0.001 |
| Recent CD4 count (cells/μl) at viraemia | ≤ 200 | 308/543 (56.7) | 1 | - | 1 | - |
|  | 201-350 | 459/693 (66.2) | 1.16 (1.07-1.27) | <0.001 | 1.13 (1.04-1.22) | 0.004 |
|  | 351-500 | 515/711 (72.4) | 1.27 (1.17-1.38) | <0.001 | 1.20 (1.11-1.29) | <0.001 |
|  | > 500 | 858/1135 (75.6) | 1.33 (1.23-1.43) | <0.001 | 1.22 (1.14-1.31) | <0.001 |
|  | Missing | 398/659 (60.4) | 1.06 (0.97-1.16) | 0.195 | 1.03 (0.95-1.13) | 0.468 |
| Data are n/N (%) unless otherwise stated. ^a^ The primary exposure effect (3-month viral suppression) is adjusted for all other variables in the table as potential confounders. ART=Antiretroviral therapy, μl=Microliter, ml=Milliliter, RR=Risk ratio, TEE=Tenofovir disoproxil fumarate plus emtricitabine plus efavirenz, TLD=Tenofovir disoproxil fumarate plus lamivudine plus dolutegravir. | | | | | | |

| **Table S 3.** Follow-up outcomes after high-level viraemia (≥1000 copies/ml) in people receiving first-line ART | | | |
| --- | --- | --- | --- |
|  | | **ART regimen at viraemia** | |
| **Variable** | **Overall**,  N = 2,163 | **TEE**,  N = 1,804 | **TLD**,  N = 359 |
| **At least one ART visit within 6 months** | 1,988 (91.9%) | 1,663 (92.2%) | 325 (90.5%) |
| **Number of ART visits within 6 months, median (IQR)** | 3 (2-4) | 3 (2-4) | 3 (2-4) |
| **Days to first ART visit within 6 months, median (IQR)** | 87 (79-105) | 87 (78-104) | 88 (83-111) |
| **Repeat 3-month viral load test done ^a^** | 1,135 (57.1%) | 945 (56.8%) | 190 (58.5%) |
| **Days to repeat 3-month viral load, median (IQR)** | 110 (85-136) | 109 (85-132) | 112 (88-146) |
| **Repeat 3-month viral load (copies/ml)** |  |  |  |
| <50 | 521 (45.9%) | 400 (42.3%) | 121 (63.7%) |
| 50-199 | 136 (12.0%) | 114 (12.1%) | 22 (11.6%) |
| 200-999 | 112 (9.9%) | 103 (10.9%) | 9 (4.7%) |
| 1000+ | 366 (32.2%) | 328 (34.7%) | 38 (20.0%) |
| **Change to 2nd-line ART within 12 months after repeat 3-month viral load ≥1000 copies/ml** | 107 (29.2%) | 104 (31.7%) | 3 (7.9%) |
| **Second-line regimens changed-to** |  |  |  |
| AZT/XTC/LPV/r | 34 (31.8%) | 32 (30.8%) | 2 (66.7%) |
| AZT/XTC/DTG | 37 (34.6%) | 37 (35.6%) | 0 (0.0%) |
| TLD | 31 (29.0%) | 31 (29.8%) | 0 (0.0%) |
| Other | 5 (4.7%) | 4 (3.8%) | 1 (33.3%) |
| **Follow-up outcome at 12 months** |  |  |  |
| Lost to follow-up | 564 (26.1%) | 486 (26.9%) | 78 (21.7%) |
| Died | 27 (1.2%) | 24 (1.3%) | 3 (0.8%) |
| Transferred out to another clinic | 172 (8.0%) | 152 (8.4%) | 20 (5.6%) |
| Retained in care | 1,400 (64.7%) | 1,142 (63.3%) | 258 (71.9%) |
| **12-month viral load done (of participants in care)** | 1,151 (82.2%) | 950 (83.2%) | 201 (77.9%) |
| **Days to 12-month viral load, median (IQR)** | 353 (283-388) | 353 (287-390) | 351 (260-380) |
| **12-month viral load (copies/ml)** |  |  |  |
| <50 | 739 (64.2%) | 585 (61.6%) | 154 (76.6%) |
| 50-199 | 101 (8.8%) | 83 (8.7%) | 18 (9.0%) |
| 200-999 | 83 (7.2%) | 77 (8.1%) | 6 (3.0%) |
| 1000+ | 228 (19.8%) | 205 (21.6%) | 23 (11.4%) |
| Data are n (%) or median (IQR). Percentages may not add up to 100 because of rounding. All percentages were calculated with the total number in the respective column headers as the denominators except otherwise stated.  ^a^ Window 1-6 months. ART=Antiretroviral therapy, AZT=Zidovudine, DTG=Dolutegravir, IQR=Interquartile range, LPV/r=Ritonavir-boosted lopinavir, μl=Microliter, ml=Milliliter, TEE=Tenofovir disoproxil fumarate plus emtricitabine plus efavirenz, TLD=Tenofovir disoproxil fumarate plus lamivudine plus dolutegravir, XTC=Emtricitabine or Lamivudine. | | | |

| **Table S 4**. Univariable and multivariable Poisson regression models of factors associated with 3-month viral suppression after high-level viraemia (≥1000 copies/ml) in people receiving first-line ART (N=1135) | | | | | | |
| --- | --- | --- | --- | --- | --- | --- |
| Variable | Level | 3-month viral suppression  (<50 copies/ml)  n/N (%) | Unadjusted RR  (95% CI) | P value | Adjusted RR  (95% CI) | P value |
| First-line regimen at viraemia | TEE | 400/945 (42.3) | 1 | - | 1 | - |
|  | TLD | 121/190 (63.7) | 1.51 (1.35-1.68) | <0.001 | 1.39 (1.22-1.58) | <0.001 |
| Age at viraemia | 15-24 | 45/118 (38.1) | 1 | - | 1 | - |
|  | 25-34 | 177/427 (41.5) | 1.08 (0.81-1.44) | 0.594 | 1.13 (0.85-1.50) | 0.400 |
|  | 35-44 | 177/373 (47.5) | 1.23 (0.97-1.56) | 0.090 | 1.25 (0.98-1.59) | 0.078 |
|  | 45+ | 122/217 (56.2) | 1.46 (1.15-1.87) | 0.002 | 1.38 (1.08-1.76) | 0.010 |
| Gender | Male | 195/402 (48.5) | 1 | - | 1 | - |
|  | Female | 326/733 (44.5) | 0.92 (0.81-1.04) | 0.187 | 0.96 (0.84-1.10) | 0.543 |
| Known active tuberculosis disease at viraemia | No | 515/1115 (46.2) | 1 | - | 1 | - |
|  | Yes | 6/20 (30.0) | 0.64 (0.29-1.40) | 0.261 | 0.78 (0.39-1.59) | 0.495 |
| Time-period of viraemia | June to July 2020 | 86/270 (31.9) | 1 | - | 1 | - |
|  | August 2020 | 94/189 (49.7) | 1.55 (1.24-1.93) | <0.001 | 1.40 (1.12-1.75) | 0.004 |
|  | September 2020 | 129/260 (49.6) | 1.55 (1.27-1.90) | <0.001 | 1.41 (1.15-1.73) | 0.001 |
|  | October 2020 | 112/211 (53.1) | 1.66 (1.37-2.01) | <0.001 | 1.50 (1.25-1.81) | <0.001 |
|  | November 2020 | 100/205 (48.8) | 1.53 (1.26-1.85) | <0.001 | 1.38 (1.14-1.67) | 0.001 |
| Recent CD4 count (cells/μl) at viraemia | ≤ 200 | 78/220 (35.5) | 1 | - | 1 | - |
|  | 201-350 | 97/237 (40.9) | 1.15 (0.90-1.48) | 0.263 | 1.18 (0.93-1.51) | 0.176 |
|  | 351-500 | 102/194 (52.6) | 1.49 (1.21-1.83) | <0.001 | 1.52 (1.22-1.90) | <0.001 |
|  | > 500 | 160/271 (59.0) | 1.67 (1.38-2.02) | <0.001 | 1.70 (1.39-2.07) | <0.001 |
|  | Missing | 84/213 (39.4) | 1.11 (0.86-1.44) | 0.409 | 1.16 (0.90-1.50) | 0.250 |
| Data are n/N (%), unless otherwise stated. ^a^ The primary exposure effect (3-month viral suppression) is adjusted for all other variables in the table as potential confounders. ART=Antiretroviral therapy, μl=Microliter, ml=Milliliter, RR=Risk ratio, TEE=Tenofovir disoproxil fumarate plus emtricitabine plus efavirenz, TLD=Tenofovir disoproxil fumarate plus lamivudine plus dolutegravir. | | | | | | |

| **Table S 5.** Univariable and multivariable Poisson regression models of factors associated with retention-in-care at 12 months follow-up after high-level viraemia (≥1000 copies/ml) in people receiving first-line ART (N=2163) | | | | | | |
| --- | --- | --- | --- | --- | --- | --- |
| Variable | Level | Retention-in-care  at 12 months  n/N (%) | Unadjusted RR  (95% CI) | P value | Adjusted RR ^a^  (95% CI) | P value |
| First-line regimen at viraemia | TEE | 1142/1804 (63.3) | 1 | - | 1 | - |
|  | TLD | 258/359 (71.9) | 1.11 (1.01-1.21) | 0.025 | 1.11 (1.02-1.22) | 0.018 |
| Age at viraemia | 15-24 | 123/244 (50.4) | 1 | - | 1 | - |
|  | 25-34 | 507/850 (59.6) | 1.18 (1.05-1.32) | 0.005 | 1.19 (1.06-1.34) | 0.003 |
|  | 35-44 | 476/688 (69.2) | 1.33 (1.19-1.49) | <0.001 | 1.37 (1.23-1.53) | <0.001 |
|  | 45+ | 294/381 (77.2) | 1.47 (1.32-1.63) | <0.001 | 1.49 (1.33-1.66) | <0.001 |
| Gender | Male | 489/763 (64.1) | 1 | - | 1 | - |
|  | Female | 911/1400 (65.1) | 1.04 (0.98-1.11) | 0.222 | 1.11 (1.03-1.20) | 0.005 |
| Known active tuberculosis disease at viraemia | No | 1375/2119 (64.9) | 1 | - | 1 | - |
|  | Yes | 25/44 (56.8) | 0.85 (0.67-1.07) | 0.155 | 0.88 (0.69-1.12) | 0.306 |
| Time-period of viraemia | June to July 2020 | 340/556 (61.2) | 1 | - | 1 | - |
|  | August 2020 | 243/348 (69.8) | 1.12 (1.01-1.24) | 0.025 | 1.08 (0.98-1.20) | 0.136 |
|  | September 2020 | 321/458 (70.1) | 1.15 (1.05-1.25) | 0.002 | 1.10 (1.01-1.20) | 0.027 |
|  | October 2020 | 277/425 (65.2) | 1.06 (0.98-1.15) | 0.151 | 1.02 (0.94-1.11) | 0.614 |
|  | November 2020 | 219/376 (58.2) | 0.95 (0.86-1.06) | 0.385 | 0.93 (0.84-1.03) | 0.159 |
| Recent CD4 count (cells/μl) at viraemia | ≤ 200 | 280/447 (62.6) | 1 | - | 1 | - |
|  | 201-350 | 268/431 (62.2) | 1.00 (0.90-1.11) | 0.999 | 0.99 (0.90-1.09) | 0.874 |
|  | 351-500 | 236/370 (63.8) | 1.02 (0.92-1.13) | 0.683 | 1.02 (0.92-1.13) | 0.722 |
|  | > 500 | 376/536 (70.1) | 1.13 (1.04-1.22) | 0.005 | 1.09 (1.01-1.18) | 0.034 |
|  | Missing | 240/379 (63.3) | 1.01 (0.91-1.11) | 0.894 | 1.03 (0.94-1.14) | 0.520 |
| Data are n/N (%), unless otherwise stated. ^a^ The primary exposure effect (retention-in-care at 12 months) is adjusted for all other variables in the table as potential confounders. ART=Antiretroviral therapy, μl=Microliter, ml=Milliliter, RR=Risk ratio, TEE=Tenofovir disoproxil fumarate plus emtricitabine plus efavirenz, TLD=Tenofovir disoproxil fumarate plus lamivudine plus dolutegravir. | | | | | | |

| **Table S 6.** Univariable and multivariable Poisson regression models of factors associated with viral suppression at 12 months follow-up after high-level viraemia (≥1000 copies/ml) in people receiving first-line ART who were retained in care at 12 months follow-up and had viral load done (N=1151) | | | | | | |
| --- | --- | --- | --- | --- | --- | --- |
| Variable | Level | Viral suppression  (<50 copies/ml)  at 12 months  n/N (%) | Unadjusted RR  (95% CI) | P value | Adjusted RR  (95% CI) | P value |
| First-line regimen at viraemia | TEE | 585/950 (61.6) | 1 | - | 1 | - |
|  | TLD | 154/201 (76.6) | 1.25 (1.14-1.38) | <0.001 | 1.24 (1.12-1.36) | <0.001 |
| Age at viraemia | 15-24 | 53/107 (49.5) | 1 | - | 1 | - |
|  | 25-34 | 264/413 (63.9) | 1.29 (1.03-1.61) | 0.027 | 1.30 (1.05-1.62) | 0.016 |
|  | 35-44 | 250/390 (64.1) | 1.29 (1.03-1.62) | 0.029 | 1.28 (1.02-1.59) | 0.029 |
|  | 45+ | 172/241 (71.4) | 1.44 (1.14-1.81) | 0.002 | 1.37 (1.08-1.72) | 0.008 |
| Gender | Male | 270/395 (68.4) | 1 | - | 1 | - |
|  | Female | 469/756 (62.0) | 0.91 (0.84-0.98) | 0.019 | 0.93 (0.85-1.02) | 0.144 |
| Known active tuberculosis disease at viraemia | No | 729/1130 (64.5) | 1 | - | 1 | - |
|  | Yes | 10/21 (47.6) | 0.74 (0.48-1.14) | 0.172 | 0.81 (0.54-1.22) | 0.317 |
| Time-period of viraemia | June to July 2020 | 181/296 (61.1) | 1 | - | 1 | - |
|  | August 2020 | 147/213 (69.0) | 1.13 (0.98-1.30) | 0.087 | 1.06 (0.93-1.21) | 0.353 |
|  | September 2020 | 188/267 (70.4) | 1.15 (1.01-1.31) | 0.034 | 1.07 (0.94-1.21) | 0.292 |
|  | October 2020 | 131/213 (61.5) | 1.00 (0.88-1.14) | 0.992 | 0.95 (0.84-1.07) | 0.410 |
|  | November 2020 | 92/162 (56.8) | 0.93 (0.77-1.11) | 0.414 | 0.88 (0.73-1.06) | 0.178 |
| Recent CD4 count (cells/μl) at viraemia | ≤ 200 | 118/222 (53.2) | 1 | - | 1 | - |
|  | 201-350 | 135/220 (61.4) | 1.16 (1.01-1.34) | 0.038 | 1.17 (1.02-1.35) | 0.030 |
|  | 351-500 | 135/204 (66.2) | 1.25 (1.10-1.42) | 0.001 | 1.25 (1.10-1.44) | 0.001 |
|  | > 500 | 233/316 (73.7) | 1.40 (1.23-1.59) | <0.001 | 1.41 (1.24-1.60) | <0.001 |
|  | Missing | 118/189 (62.4) | 1.18 (1.02-1.35) | 0.022 | 1.21 (1.06-1.39) | 0.006 |
| Data are n/N (%), unless otherwise stated. ^a^ The primary exposure effect (viral suppression at 12 months) is adjusted for all other variables in the table as potential confounders. ART=Antiretroviral therapy, μl=Microliter, ml=Milliliter, RR=Risk ratio, TEE=Tenofovir disoproxil fumarate plus emtricitabine plus efavirenz, TLD=Tenofovir disoproxil fumarate plus lamivudine plus dolutegravir. | | | | | | |
